# Multivariate Associations Between Structural Brain Networks, Genetics, Environments, and Cognitive-Psychopathological Traits in Children

**DOI:** 10.1101/2023.07.24.23293075

**Authors:** Jungwoo Seo, Eunji Lee, Bo-Gyeom Kim, Gakyung Kim, Yoonjung Yoonie Joo, Jiook Cha

## Abstract

Brain development in childhood is shaped by complex interactions between genetic predispositions, environmental influences, and neural connectivity, yet how these factors jointly contribute to cognitive and mental health outcomes remains unclear. Structural brain networks, quantified through graph-theoretic measures, have been linked to cognition and psychiatric risk, but the extent to which genetic architecture and environmental exposures shape these networks, and whether brain networks mediate these influences, is not well understood. Here we show that genetic predispositions related to cognitive ability and socioeconomic status (SES) exhibit the strongest covariation with structural brain network topology in children. Using sparse canonical correlation analysis (SCCA) on ABCD Study data (**N** = 10,343), we identified robust associations between brain network properties, polygenic scores for cognition, SES indicators, and cognitive-psychopathological phenotypes. Mediation analysis further demonstrated that structural brain networks partially mediate the influence of genetic and environmental factors on cognitive performance and mental health outcomes, suggesting that neurodevelopmental trajectories may be shaped by both genetic liability and modifiable environmental conditions. These findings provide empirical support for a multivariate, systems-level perspective on brain development and cognitive-psychopathological variation in youth. By elucidating shared neural substrates underlying genetic and environmental influences, this work advances our understanding of brain network development and highlights potential pathways for individualized interventions and predictive modeling in developmental psychiatry and neuroscience.

## Introduction

Childhood and adolescence are critical periods for brain development (Bethlehem et al., 2022). Proper brain development during this time is vital for cognitive and behavioral maturation (Bunge & Wright, 2007; Luna et al., 2010) and mental health (Fornito et al., 2015; Paus et al., 2008). This development is shaped by a complex interplay between genetic predispositions, environmental influences, and brain network dynamics, yet the precise mechanisms underlying these relationships remain poorly understood. Therefore, understanding the connections between the brain, cognitive-behavioral traits in children, and the impact of genetics and the environment on brain development is crucial in developmental and clinical neuroscience.

The Adolescent Brain and Cognitive Development (ABCD) study provides an unprecedented opportunity to examine these interrelations in a large, representative sample (Jernigan et al., 2018). Leveraging multimodal neuroimaging, genome-wide polygenic scores (PGSs), and extensive environmental and behavioral assessments, the ABCD dataset enables a more comprehensive modeling of developmental trajectories than previously possible.

The brain is best understood as a complex, interconnected network, rather than a collection of isolated regions. Graph-theoretic approaches to brain connectivity provide powerful tools to quantify structural network properties such as integration (e.g., global efficiency), segregation (e.g., modularity), and centrality (e.g., degree centrality) (Rubinov & Sporns, 2010). Diffusion MRI and tractography allow for the reconstruction of white matter networks, enabling researchers to examine how these structural connections support cognition (Jeurissen et al., 2019; Sotiropoulos & Zalesky, 2019). Notably, individual differences in brain network organization have been linked to cognitive abilities (Bathelt, Gathercole, Butterfield, et al., 2018; Kim et al., 2016; Koenis et al., 2015; Ma et al., 2017; Suprano et al., 2020), psychiatric risk (Alexander-Bloch et al., 2010; Collin et al., 2017; Rudie et al., 2012), and environmental influences such as socioeconomic status (Kim et al., 2019; Tooley et al., 2020). However, how genetic and environmental factors jointly shape structural brain networks, and whether these networks mediate their influence on cognitive and psychiatric outcomes, remains poorly understood.

Twin and heritability studies suggest that brain network properties are, at least in part, genetically influenced, with estimates of heritability for measures like global efficiency ranging from 25% to 70% (Koenis et al., 2015; van den Heuvel et al., 2013). Yet, the specific genetic variants shaping these structural networks—particularly in the context of cognitive and psychiatric outcomes—remain poorly characterized. Investigating this gene-brain-behavior interplay is essential for understanding how biological predispositions manifest in cognitive and mental health outcomes during development.

Despite valuable insights from previous studies, most prior research has employed univariate approaches, limiting the ability to capture the multivariate complexity of the gene/environment-brain-behavior relationship. To address this limitation, canonical correlation analysis (CCA) is useful for investigating holistic relationships underlying a set of variables. CCA models multivariate associations linking sets of variables from two or more domains by maximizing the canonical correlation between them (Hotelling, 1936; Wang et al., 2020). Indeed, CCA has been widely applied in studying links between brain connectivity, cognitive function, genetics, and environmental factors (Alnaes et al., 2020; Fernandez-Cabello et al., 2022; Modabbernia et al., 2021; Smith et al., 2015; Wang et al., 2020). However, traditional CCA methods may struggle with high-dimensional datasets, necessitating more advanced approaches. To further address these limitations, we apply sparse canonical correlation analysis (SCCA) (Witten et al., 2009), a multivariate method that identifies maximally covarying patterns between genetic factors, brain network properties, and behavioral traits while reducing dimensionality and enhancing interpretability.

Utilizing the ABCD dataset’s expansive genetic-environmental-neuroimaging-behavioral data, this study aims to answer two fundamental questions. Firstly, “How do genetic predispositions/environmental influences shape structural brain network properties in children?”; Secondly, “Do brain network properties mediate the influence of genetic and environmental factors on cognitive or psychiatric phenotypes?”

By integrating genetics, environmental, neuroimaging, and behavioral assessments in a large, developmentally significant cohort, this study seeks to elucidate the complex gene-environmental-brain-behavior pathways that shape cognitive and mental health outcomes. This research contributes to a more comprehensive understanding of the biological and environmental foundations of brain development and lays the groundwork for future interventions aimed at optimizing cognitive and psychological well-being in youth.

## Methods and Materials

### ABCD participants

We used genetic, neuroimaging, environmental, and phenotypic data from the Adolescent Brain Cognitive Development (ABCD) study (http://abcdstudy.org), specifically from release 2.0 for genetic and neuroimaging data, release 3.0 for genetic ancestry information, and release 5.1 for environmental and phenotypic data. The ABCD study, the largest longitudinal investigation of brain development and child health in the United States, recruited multiethnic children (N=11,875) aged 9-10 years from 21 research sites. The self-reported ethnicities of participants included 52.3% White, 20.3% Hispanic, 14.7% Black, and 12.5% Asian and others. All participants provided informed assent, and their parents or legal guardians provided informed consent before participating in the study.

### Genotype Data and Polygenic Scores

The genotype data used in this study were obtained from the ABCD cohort, with DNA samples genotyped at Rutgers University Cell and DNA Repository (RUCDR) using the Affymetrix NIDA Smoke Screen Array. Standard quality control (QC) procedures were applied to remove variants with low genotype call rate and minor allele frequency (MAF). Genotype imputation was performed using the Michigan Imputation Server (Das et al., 2016) with the 1000 Genomes reference panel (Genomes Project et al., 2015). We further filtered out imputed variants that did not meet our QC criteria. To account for population structure and relatedness, principal component analysis (PCA) and kinship-based filtering were used to exclude close relatives (Conomos et al., 2015; Conomos et al., 2016).

Polygenic scores (PGSs) for 30 cognitive, psychiatric, and behavioral traits were calculated based on genome-wide association study (GWAS) summary statistics. These included PGSs for cognitive performance, educational attainment, schizophrenia, bipolar disorder, depression, insomnia, body mass index, and automobile speed propensity, among others. The PGSs were derived using PRS-CS (Ge et al., 2019), a Bayesian regression approach, with optimal hyperparameter selection.

The genotype data and polygenic scores used in this study are identical to those reported in (Joo et al., 2024). A complete list of the 30 polygenic scores, along with further methodological details on genotype data processing, GWAS sources, ancestry-based adjustments, and validation procedures can be found in the **Supplementary Materials** or (Joo et al., 2024).

### Environmental Factors

To investigate the relationship between children’s brain network properties and their environment, we analyzed 56 variables representing various environmental aspects. These variables included indicators of family and neighborhood socioeconomic status, such as family income, parental education level, marital status, and neighborhood deprivation based on the Area Deprivation Index (ADI). Additionally, variables related to prenatal substance exposure (e.g., tobacco, alcohol, cocaine, and marijuana), as well as parental and family factors such as parental acceptance and family conflict, were included.

To maximize statistical power, we imputed missing data to include as many samples as possible in the analysis. To account for uncertainty in imputing missing values and to improve the accuracy and reliability of the imputed values, we employed the Multiple Imputation by Chained Equations (MICE) method (Van Buuren & Groothuis-Oudshoorn, 2011). We set the number of imputation iterations to 40 to ensure stable convergence. All imputation procedures were conducted using the `statsmodels` package in Python. To ensure that our analysis was robust to imputation, we also conducted analyses using only the complete data without any missing values and included these results as supplementary material.

### Phenotype Data

To explore the relationship between children’s brain network properties and their cognitive ability, mental well-being, and physical health, we examined 86 phenotype variables. To evaluate cognitive ability, we examined the NIH Toolbox measurements, which included fluid, crystallized, and overall cognition scores, along with domain-specific task scores for episodic memory, executive function, language, and processing speed (Weintraub et al., 2013). For assessing the children’s mental health, our analysis encompassed a broad range of emotional and behavioral measurements, including instruments such as the Child Behavioral Checklist (CBCL), the Kiddie-Structured Assessment for Affective Disorders and Schizophrenia for DSM-5 (KSADS-5), and the Prodromal Psychosis Scale (PPS). We also included measures of behavioral tendencies, such as behavioral inhibition and activation (BIS/BAS), which assess avoidance behaviors, reward sensitivity, and behavioral control; impulsivity traits (UPPS), which measure dimensions such as urgency, perseverance, and sensation seeking; and sleep-related problems, which provide insights into sleep patterns, quality, and related disorders. For the children’s physical health, we included data such as physical activity. To maximize statistical power, missing values in the phenotype data were imputed using the Multiple Imputation by Chained Equations method (Van Buuren & Groothuis-Oudshoorn, 2011).

### Structural Brain Network Construction

Detailed procedures for acquiring and preprocessing MRI data are described in (Kim et al., 2022). In brief, we used structural and diffusion MRI data acquired by the ABCD study (Casey et al., 2018) from data release 2.0. The preprocessing steps, as detailed in (Kim et al., 2022), included eddy current and head motion correction, diffusion gradient adjustment, and various distortion corrections. Quality control (QC) was performed using freesurfer QC metric (fsqc_qc) and raw dMRI QC metric (iqc_dmri_1_qc_score). To estimate brain structural networks from neuroimaging, individual connectome data was generated. This was achieved by applying MRtrix3 (Tournier et al., 2019) to the preprocessed dMRI data to estimate whole-brain white matter tracts and generate individualized connectomes. Probabilistic tractography was performed using constrained-spherical deconvolution (CSD) (Calamante et al., 2010; Tournier et al., 2007) with random seeding across the brain and target streamline counts of 20 million. Initial tractograms were filtered using spherical-deconvolution informed filtering (2:1 ratio)(Smith et al., 2013), resulting in a final streamline count of 10 million. An 84x84 whole-brain connectome matrix was generated for each participant using the T1-based parcellation and segmentation from FreeSurfer with Desikan-Killiany atlas (Desikan et al., 2006)(68 nodes for the cortical regions and 16 nodes for the subcortical regions). This approach ensured that individual participants’ connectomes were based on their neuroanatomy. The computation was conducted on supercomputers at Argonne Leadership Computing Facility Theta and Texas Advanced Computing Center Stampede2.

### Brain Network Measures (BNMs)

We used the connectome matrix to construct an undirect weighted graph representing the structural brain network. Nodes and edges in the graph represent parcellated gray matter regions and connections between them, respectively. Connection strength was quantified by the streamline counts. To account for the potential false positive connections generated by probabilistic tractography and their impact on network topology, we eliminated extremely weak connections (streamline counts less than 3). After thresholding, we excluded individuals with at least one isolated node, assuming all brain regions are communicable via at least one path.

We calculated 13 different types of brain network measurements (BNMs) representing different aspects of brain network’s property (Rubinov & Sporns, 2010; van den Heuvel & Sporns, 2011). We calculated eight global graph metrics (including network density, modularity, normalized modularity, normalized average clustering coefficient, normalized characteristic path length, global efficiency, normalized global efficiency, small worldness) and five nodal graph metrics (including degree, strength, clustering coefficient, betweenness centrality, nodal efficiency) to represent brain network’s global and regional properties. All graph measures were calculated using the package Brain Connectivity Toolbox (https://sites.google.com/site/bctnet/).

### Statistical analysis

#### Sparse Canonical Correlation Analysis

To examine a latent mode of covariation between structural brain network properties and various polygenic scores, environmental factors, and phenotypic outcomes, we used sparse canonical correlation analysis (Witten et al., 2009) between brain network measures and three types of non-imaging data (i.e., PGSs, environmental variables, phenotype variables) separately. While traditional CCA can be effective, it often suffers from overfitting and interpretability issues in high-dimensional datasets. SCCA, with its L1 regularization, addresses these issues by producing sparse solutions that enhance interpretability and reduce overfitting. Although our dataset is not high-dimensional enough to make traditional CCA infeasible, we chose SCCA to identify interpretable patterns and ensure robust results.

The most popular algorithm for sparse canonical correlation analysis is penalized matrix decomposition (PMD)(Witten et al., 2009), which solves optimization problem of below equation for given two sets of data matrix *X_n×p_*, *Y_n×q_* (n: sample size; p, q: the number of variables of domain X and Y respectively; u, v: canonical weights of domain X and Y respectively; c1, c2: regularization parameter)

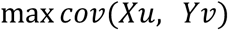

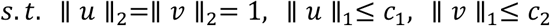

To interpret Witten’s sparse canonical correlation analysis as correlation maximization, we need to assume covariance matrices *X^T^X*, *Y^T^Y* are identity matrices (Witten et al., 2009). But in our study with the high dimensional brain datasets, the assumption is hardly satisfied. For this reason, we interpreted Witten’s sparse canonical correlation analysis as a maximizing covariance algorithm between two sets of variables rather than maximizing correlation.

To test the generalizability of the sparse canonical correlation analysis results, we split the dataset into a training and test set. To reduce ABCD site-sensitive bias, we performed a stratified train (80%) - test (20%) split based on site ID. For the sparse canonical correlation analysis with PGS and brain network measures, we only used participants classified as genetically European ancestry to control genetic confounding effects as the main analysis. To ensure that these analyses generalize to a multiethnic dataset, we conducted additional analyses on the multiethnic dataset. In the multiethnic analysis, genetic ancestry was included as an additional covariate. **Table 1 and Supplementary Table 1** summarize the demographic information of the samples included in main and supplementary analysis, respectively.

**Table 1.**
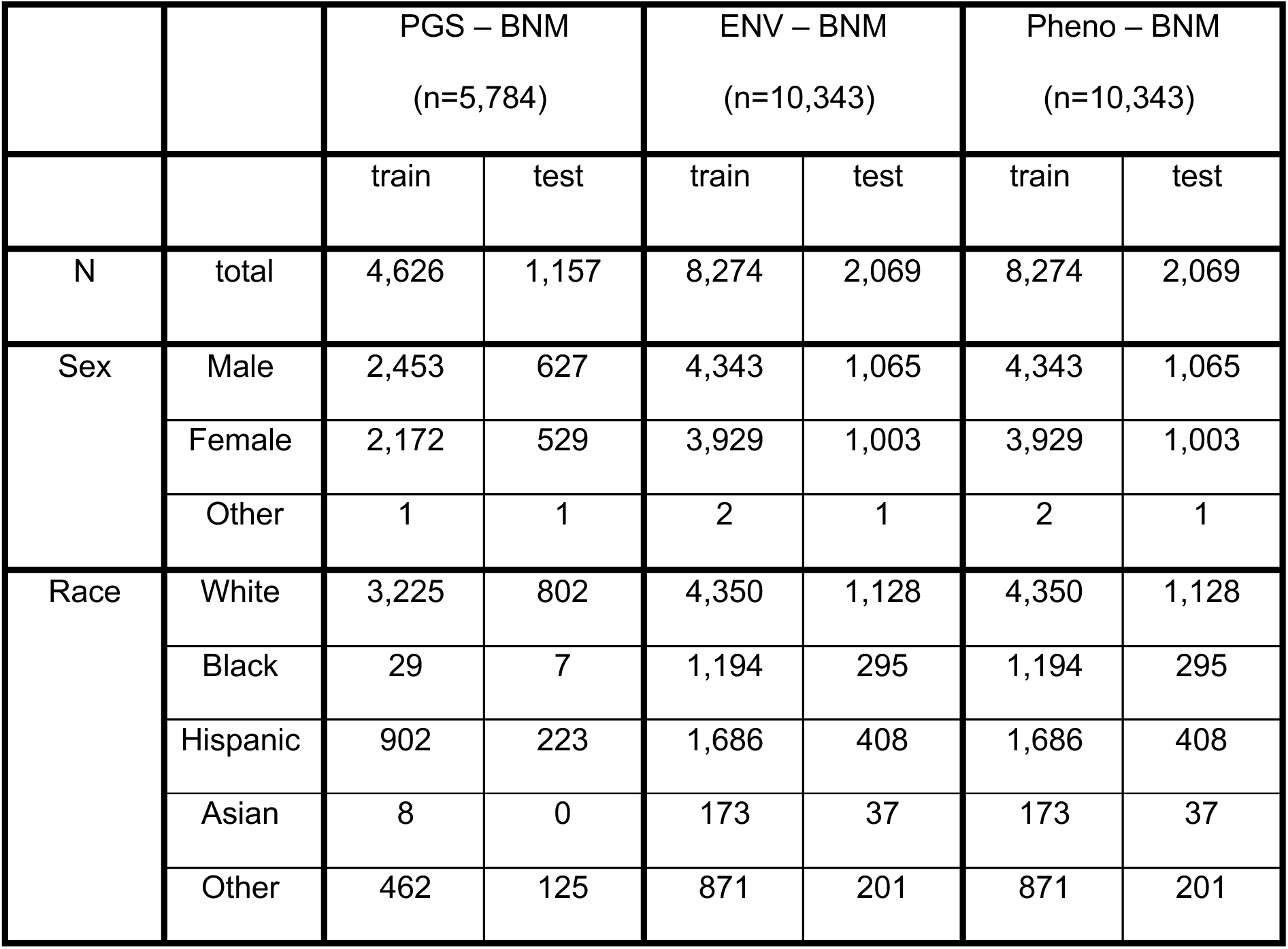
Demographic information of the main analysis participants.

We attempted to control for potential confounding effects from age, sex, self-reported race/ethnicity, ABCD study site, and handedness. Similar to (Modabbernia et al., 2021), we controlled these potential confounding effects by regressing out the variance explained by age, sex, age * sex, age^2, age^2 * sex, self-reported race/ethnicity, ABCD study site, and handedness from all variables prior to performing SCCA analysis. For binary variables such as KSADS diagnosis, we used logistic regression to regress out these effects. The residualized data was then used as input for sparse canonical correlation analysis (SCCA). We used `SCCA_PMD` function from the Python ‘cca-zoo’ package. To ensure that the main findings are robust and not driven by the selection of specific covariates, we performed additional analyses using a reduced set of covariates (age, sex, self-reported race/ethnicity, ABCD study site, and handedness).

We selected optimal L1 regularization parameters from 5-fold cross validation searching from 0.1 to 1 with a step size of 0.05 for both X and Y variables respectively. The optimal L1 parameter combination was selected to maximize the covariance of validation set between canonical variates of the first component.

For each sparse canonical correlation analysis, we extracted five modes of covariance. To examine the statistical significance of each mode, we used a permutation test. By randomly shuffling the rows of one dataset and remaining the other, we generated 5,000 permutation sets. The p-value of each component was calculated based on the number of permutation sets having greater covariance than that obtained from the original dataset, and FDR-correction was done within each CCA.

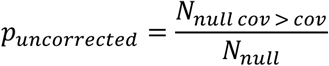

Selected variables and their loading depend on the input sample. To find variables reliably related to each mode, we used bootstrap resampling. We randomly resampled 5,000 times with replacement and assessed the 95% confidence interval of each variable’s loading and how consistently it was selected. We interpreted the significant modes based on loading patterns of variables whose 95% confidence interval of loading does not cross zero (Xia et al., 2018) and selected more frequently than expected by chance (i.e., more frequently selected than expected by binomial distribution). Because sparse canonical correlation analysis with bootstrap sample may change the order of components (axis rotation) and signs (reflection) (Misic et al., 2016; Xia et al., 2018), the re-alignment procedure is needed to estimate confidence interval of loading properly. We matched the components and signs based on cosine similarity of weight vectors obtained from original dataset and bootstrap sample. To assess the reproducibility of the findings, we applied the model to the held-out test set and estimated significance of each mode through the permutation test.

#### Mediation Analysis

After performing sparse canonical correlation analysis (SCCA), we tested whether brain network properties mediate the influence of genetic and environmental factors on cognitive and psychiatric phenotypes. Since SCCA identifies latent modes of covariation between brain network properties and multiple domains, the SCCA-derived variates serve as optimal summary representations that capture the dominant axes of covariation among brain network properties, genetic predispositions, environmental influences, and phenotypic traits. We used these summary scores in mediation analysis to test the hypothesized gene/environment– brain network–phenotype pathway while reducing high-dimensional data into interpretable components.

We used the same covariates as in the SCCA, which include age, sex, age * sex, age^2, age^2 * sex, self-reported race/ethnicity, ABCD study site, and handedness. The two-sided p-values for each path were estimated from 500 bootstrap samples using the `mediation_analysis` function from the Python ‘pingouin’ package.

## Results

Using sparse canonical correlation analysis (SCCA), we investigated relationships between structural brain network properties and three domains: genome-wide polygenic scores (PGSs), environmental factors, and phenotypic outcomes. **Figure 1** presents only variables whose 95% confidence intervals for loadings do not cross zero, ensuring reliable positive or negative associations.

**Figure 1.**
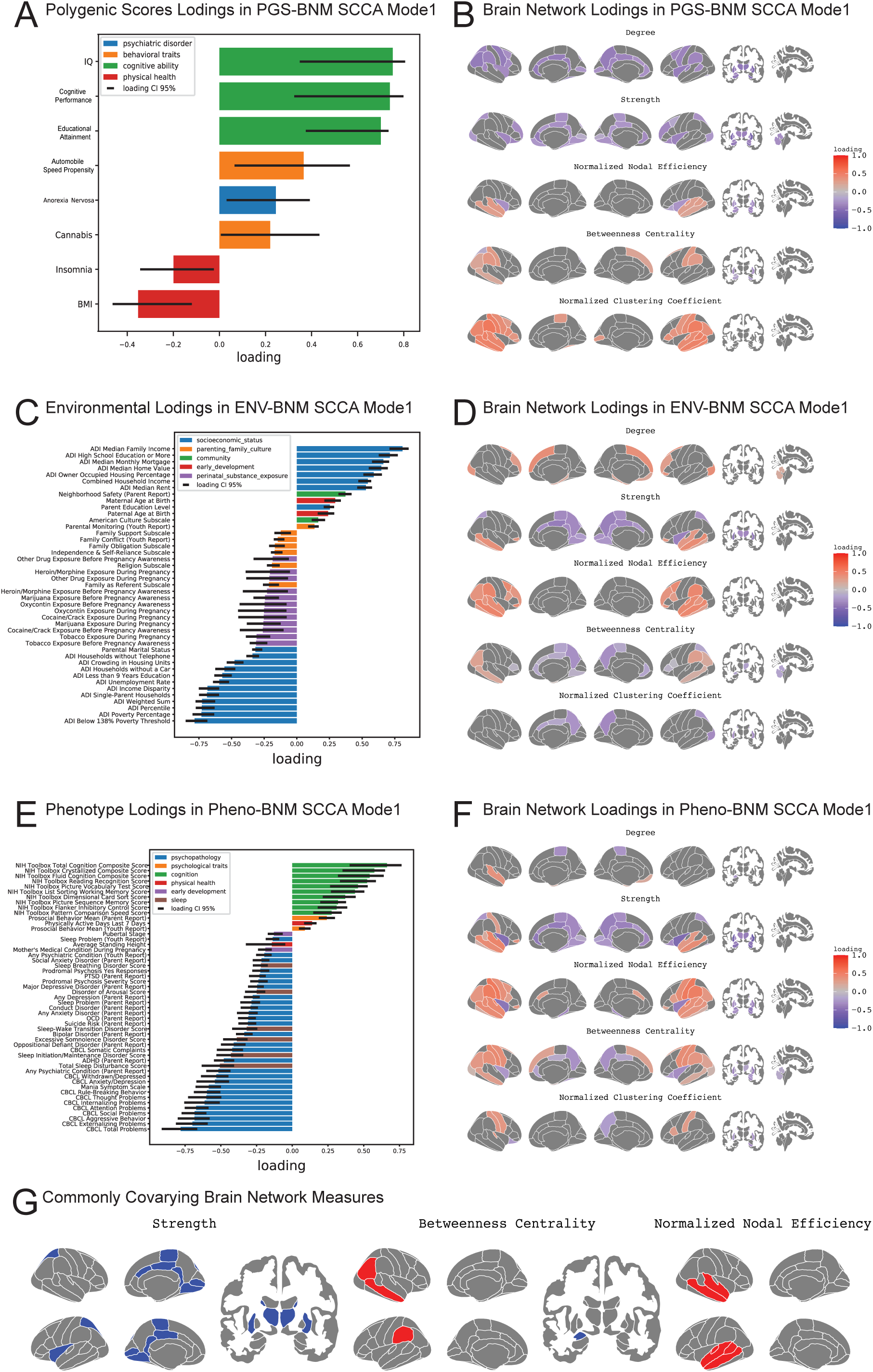
Loading patterns of principal mode of sparse canonical correlation analysis. The SCCA results for the first modes of PGS-BNM, ENV-BNM, and Pheno-BNM. Only variables with 95% confidence intervals, estimated from 5,000 bootstrap samples, that do not cross zero are shown. Results for other variables can be found in supplementary figures. (A) The loadings of significant PGS variables in the PGS-BNM mode 1. The error bars represent the 95% confidence interval of the loading, estimated from the 5,000 bootstrap samples. The color of each bar represents the category to which the variable belongs. (B) The loadings of significant nodal brain network measures in the PGS-BNM mode 1. The loading patterns were visualized with R-package ‘*ggseg’* (Mowinckel & Vidal-Pineiro, 2020). (C) The loadings of significant environmental variables in the ENV-BNM mode 1. (D) The loadings of significant nodal brain network measures in the ENV-BNM mode 1. (E) The loadings of significant phenotype variables in the Pheno-BNM mode 1. (F) The loadings of significant nodal brain network measures in the Pheno-BNM mode 1. (G) Brain network measures that their loadings are commonly significant in PGS-BNM mode 1, ENV-BNM mode1, and Pheno-BNM mode1.

### Covariation between polygenic scores and structural brain network properties

Our analysis identified a statistically significant mode of covariation between 30 genome-wide polygenic scores and brain network measures (PGS-BNM mode 1, *p* = 0.001, *cov* = 0.895, *r* = 0.143, FDR corrected).

In this mode of PGSs **(Figure 1A)**, polygenic scores reflecting cognitive ability (e.g., IQ and educational attainment) showed the strongest positive loadings, while the polygenic score for BMI showed a negative loading. Additionally, polygenic scores for automobile speed propensity and cannabis use had moderate positive loadings, whereas those for insomnia showed moderate negative loadings. Given these dominant loading patterns, this mode represents variance along the ‘Cognitive-Obesity Genetic Axis,’ primarily reflecting genetic traits related to cognitive ability. Loadings for polygenic scores not shown in **(Figure 1A)** are provided in **(Supplementary Figure 1)**.

Higher values in this mode were linked to increased nodal efficiency in the temporal gyri, betweenness centrality in the supramarginal and post-central gyri, and higher clustering coefficients in the temporal, parietal, central, and inferior frontal gyri **(Figure 1B)**. Conversely, higher values in this mode were associated with lower connectivity in the cingulate cortex, insula, and subcortical regions, along with reduced nodal efficiency in the insula and subcortical regions. Regarding global brain network measures, density (-0.613) and global efficiency (-0.421) showed negative loadings, while the normalized average clustering coefficient (0.542) showed a positive loading.

Results from the multi-ethnic analysis were consistent with those observed in the European-only analysis **(Supplementary Figure 2)**. When we controlled for confounding effects using a simplified set of covariates, the results remained largely unchanged. However, when the SCCA model trained on the training set was applied to the test set, the p-value was marginally significant (p = 0.0534).

### Covariation between environmental factors and structural brain network properties

Among the five modes analyzed, only the first mode was statistically significant and generalized to the hold-out test set (ENV-BNM mode 1, p < 0.001, cov = 1.251, cor = 0.146, FDR corrected). The environmental component of this mode predominantly captured variance related to socioeconomic status (SES) **(Figure 1C)**.

Notably, neighborhood-level SES variables, such as ADI median family income and ADI education level, showed the strongest positive loadings. Similarly, family-level SES variables, including household income, also showed positive associations, though their loadings were slightly lower than those of neighborhood-level SES measures. Variables reflecting neighborhood-level socioeconomic deprivation, such as ADI poverty indices, showed strong negative loadings. Additionally, prenatal substance exposures—such as tobacco exposure during pregnancy—showed moderate negative associations with this component. These results suggest that this component reflects a spectrum of socioeconomic status from socioeconomic advantage to disadvantage.

On the brain network side **(Figure 1D)**, connection strength and nodal efficiency in the temporal gyrus showed positive loadings, and connection strength, betweenness centrality, and clustering coefficient in regions such as the cingulate cortex and precuneus showed negative loadings. Regarding global brain network measures, normalized global efficiency (0.360) showed positive loading.

These results were robust across different sets of covariates. When missing values were handled by dropping incomplete cases rather than through imputation, the environmental component remained consistent. The general loading patterns of brain network properties were also similar; but, the overall magnitude of the loadings was slightly lower than in the main analysis, possibly due to reduced statistical power **(Supplementary Figure 4)**.

### Covariation between phenotypes and structural brain network properties

Among the five modes of covariation identified by SCCA, the first two modes were statistically significant and generalized to the hold-out test sets (Pheno-BNM mode 1: p < 0.001, cov = 1.421, r = 0.143; Pheno-BNM mode 2: p < 0.001, cov = 0.923, r = 0.123, all p-values were FDR corrected).

The first mode captured covariation between brain network properties and an integrated measure of cognitive ability and psychopathology in children **(Figure 1E)**. Cognitive ability-related scores (e.g., NIH Toolbox scores) showed positive loadings, while psychopathology-related scores (e.g., CBCL scores) showed negative loadings. These loading patterns suggest that the first phenotype mode reflects variation along the cognitive ability-psychopathology axis.

Brain network properties associated with this mode showed positive loadings for nodal efficiency and betweenness centrality in the temporal and parietal cortices. In contrast, connection strength in the insula, cingulate, precuneus, and subcortical regions showed negative loadings. Additionally, nodal efficiency in the insula and subcortical regions was negatively associated with this mode **(Figure 1F)**. No significant global brain network measures were identified for this mode.

In phenotype mode 2 **(Figure 2)**, positive loadings were observed for standing height, being born prematurely, having hearing or vision issues, and experiencing obstetric complications. Although the uncertainty in loading estimation is considerable, abnormal behavior (CBCL scores) also exhibited the highest positive loading in this mode **(Supplementary Figure 7)**. On the brain network properties side, lower connection strength and nodal efficiency were observed in the precentral and postcentral gyri, superior frontal gyrus, and thalamus. Degree in the middle temporal gyrus, superior frontal gyrus, and postcentral gyrus also tended to be lower. Among global brain network metrics, normalized modularity (-0.195), global efficiency (-0.343), and normalized global efficiency (-0.487) showed negative loadings, while modularity (0.534) showed positive loading.

**Figure 2.**
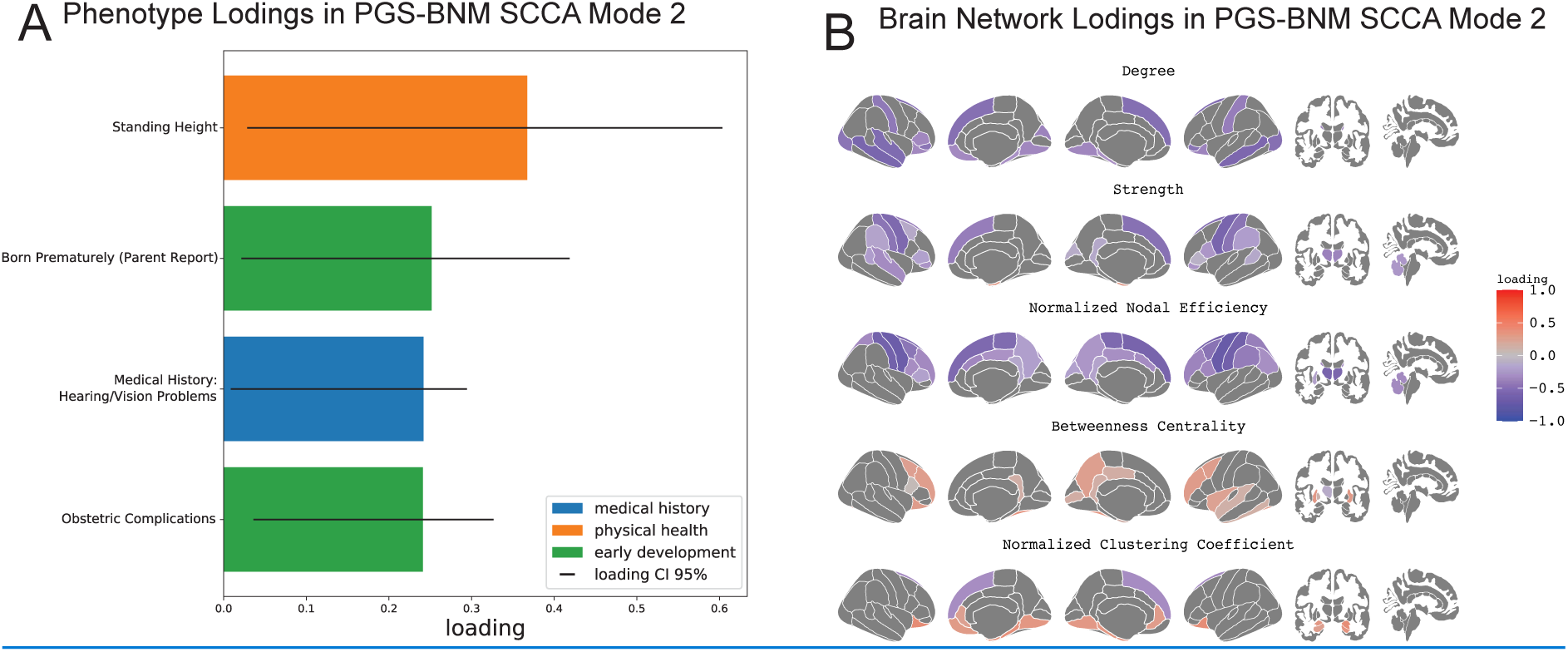
Loading patterns of second mode of Pheno-BNM sparse canonical correlation analysis. (A) The loadings of significant phenotype variables in the Pheno-BNM SCCA mode 2. (B) The loadings of significant nodal brain network measures in the Pheno-BNM SCCA mode 2.

The findings remained stable across different covariate sets. When missing values were handled by excluding incomplete cases instead of using imputation, the mode 1 results remained consistent **(Supplementary Figure 6)**. However, in mode 2, unlike the imputed case, only height, weight, and BMI exhibited reliable positive loadings, while CBCL, despite its high estimation uncertainty, showed a strong negative loading **(Supplementary Figure 8).**

### Shared Covariation Patterns in Brain Network Properties

Our SCCA identified certain brain network properties that showed similar covariation patterns across genetic factors related to cognitive ability, socioeconomic status, and phenotypes of cognitive ability-psychopathology. **(Figure 1G)** presents the brain network properties that shared common loading patterns with the primary SCCA modes for polygenic scores (PGS mode 1), environmental factors (ENV mode 1), and phenotypes (Pheno mode 1). Across these domains, brain network measures such as nodal efficiency in the temporal gyrus consistently showed positive loadings, while measures like connection strength in the posterior cingulate and subcortical regions consistently showed negative loadings.

### Mediation Analysis

The observed covariation patterns in brain network properties across genetic, environmental, and phenotypic domains led us to hypothesize that structural brain networks mediate two key relationships: (1) between genetic factors related to cognitive ability and the cognitive ability-psychopathology phenotype, and (2) between socioeconomic status and the cognitive ability-psychopathology phenotype. To test these hypotheses, we conducted mediation analyses using SCCA-derived summary scores from each domain.

Our analysis revealed significant mediation effects in both cases. Structural brain network properties partially mediated the relationship between polygenic scores for cognitive ability and the cognitive ability-psychopathology phenotype (indirect effect = 0.023, p < 0.001; **Figure 3A**), as well as between socioeconomic status and the cognitive ability-psychopathology phenotype (indirect effect = 0.015, p < 0.001; **Figure 3B)**.

**Figure 3.**
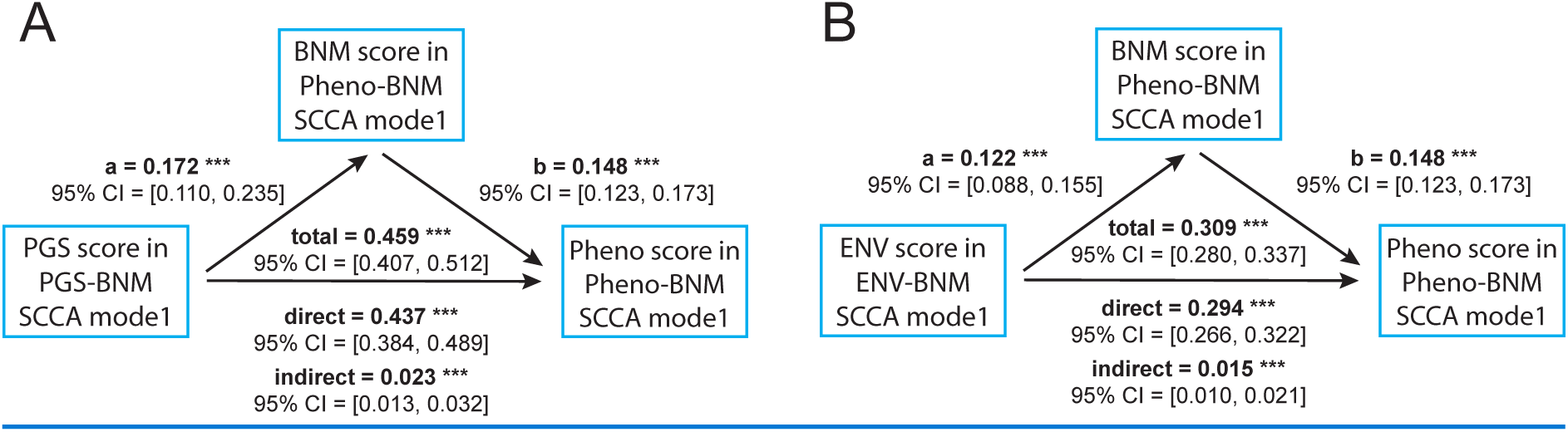
Results of mediation analysis investigating the gene-brain network-phenotype and environment-brain network-phenotype pathways. Mediation analysis was conducted using canonical variates derived from SCCA for polygenic scores, brain network measures, phenotypes, and environmental factors. (A) Brain network properties mediating the relationship between polygenic scores (reflecting cognitive ability) and phenotype scores (reflecting cognitive ability-psychopathological traits). (B) Brain network properties mediating the relationship between environmental factors (reflecting socioeconomic status) and phenotype scores (cognitive ability-psychopathological traits).

## Discussion

This study investigated how structural brain network properties covary with genetic, environmental, and phenotypic factors in 9–10-year-old children and whether these properties mediate genetic and environmental effects on cognitive-behavioral outcomes. By integrating polygenic scores (PGS), environmental variables, and cognitive-behavioral traits using sparse canonical correlation analysis (SCCA), we identified shared covariation patterns in brain network properties across these domains. Previous studies have demonstrated that structural brain network properties are heritable (Koenis et al., 2015; van den Heuvel et al., 2013), but the specific genetic contributions to brain network organization in children remain underexplored. Our findings address this gap by providing evidence that cognitive ability-related genetic factors, socioeconomic status, and cognitive-psychopathological phenotypes are key determinants of structural brain network variations in preadolescents. Moreover, mediation analyses reveal that structural brain network properties serve as intermediaries between genetic/environmental influences and cognitive-psychopathological outcomes. This suggests that variations in brain network organization may provide a mechanistic pathway through which early-life genetic and environmental factors contribute to individual differences in cognitive and mental health outcomes.

Our results reveal that structural brain network properties exhibit distinct yet overlapping covariation patterns with genetic, environmental, and phenotypic factors. Specifically, brain network measures such as nodal efficiency in the temporal and parietal cortices were consistently associated with cognitive ability-related genetic factors, higher socioeconomic status, and better cognitive performance. Conversely, weaker connectivity in the posterior cingulate, insula, and subcortical regions was commonly linked to genetic risk for lower cognitive ability, socioeconomic disadvantage, and increased psychopathology. Consistent with prior studies (Alnaes et al., 2020; Fernandez-Cabello et al., 2022; Modabbernia et al., 2021; Smith et al., 2015), these findings suggest that children’s brain network properties also covary along spectrums of ‘positive-negative’ factors across genetic, environmental, and phenotypic dimensions. Unlike previous studies that explored covariation between brain features and only some of these domains, our study examined all three factors simultaneously, identifying a common set of brain network properties that covary across genetic, environmental, and phenotypic factors. These findings highlight the potential of these brain networks as key substrates underlying cognitive and mental health disparities, shaped by both genetic predisposition and environmental influences during childhood.

Our findings align with established neurodevelopmental models, which suggest that childhood and adolescence are characterized by a shift from subcortical-driven to cortico-cortical-dominated network organization (Baker et al., 2015; Langen et al., 2018; Menon, 2013; Sato et al., 2015). Specifically, favorable genetic and phenotypic traits related to cognitive ability and SES were associated with greater nodal efficiency and betweenness centrality in the temporal and parietal cortices and reduced connectivity in the posterior cingulate, insula, and subcortical regions. These patterns are consistent with established neurodevelopmental trajectories, which involve a progressive strengthening of long-range cortico-cortical connectivity (Hwang et al., 2013; Oldham & Fornito, 2019) and reduction in subcortical connectivity as higher-order networks become more specialized (Baker et al., 2015; Langen et al., 2018; Sato et al., 2015). Interestingly, the associations between genetic, environmental, and phenotypic factors and brain network organization mirror expected patterns of cortical-subcortical reorganization, wherein cortico-cortical integration strengthens while subcortical connectivity decreases. These findings suggest that genetic and environmental influences may shape the timing or pace of this cortical-subcortical transition, potentially accelerating or delaying neurodevelopmental trajectories and influencing individual differences in cognitive and mental health outcomes (Heller et al., 2016).

Children with higher cognitive performance tend to have greater nodal efficiency and betweenness centrality in the temporal, parietal, and superior frontal regions—areas crucial for higher-order cognitive functions such as language, semantic processing, abstract reasoning, and working memory (Binder et al., 2009; Culham & Kanwisher, 2001; du Boisgueheneuc et al., 2006; Price, 2012; Visser et al., 2012). This suggests that enhanced network integrity in these regions supports cognitive ability by efficient communication across the brain. Consistent with this, our results support the parieto-frontal integration theory (P-FIT) of intelligence (Basten et al., 2015; Jung & Haier, 2007), which posits that effective information integration across distributed networks, particularly involving the frontal, parietal, and temporal regions, underlies intelligence. Notably, while previous studies have linked global efficiency with cognitive performance (Bathelt, Gathercole, Butterfield, et al., 2018; Kim et al., 2016; Ma et al., 2017), our findings—derived from a largest to date sample—suggest that regional network efficiency, rather than global efficiency, is more strongly associated with cognitive ability. This highlights the importance of region-specific network topologies in understanding neurodevelopmental differences.

Although neighborhood-level socioeconomic status (SES) and family-level SES are both associated with children’s brain development, neighborhood-level SES has a unique relationship with brain structure and functional networks, distinct from that of family-level SES (Rakesh et al., 2022; Tooley et al., 2020). In our results, the link between neighborhood-level SES and structural brain network properties was more pronounced than associations with family-level SES.

Prenatal substance exposure, particularly to tobacco and marijuana, exhibited moderate negative loadings within the SES-related brain network mode, reinforcing prior findings that lower socioeconomic status is associated with increased prenatal exposure to neurotoxic substances (Gu et al., 2024; Metz et al., 2018; Mravcik et al., 2020). This association likely reflects a complex interplay between socioeconomic adversity and prenatal environmental stressors, both of which have been implicated in shaping neurodevelopmental trajectories (El Marroun et al., 2016; Ross et al., 2015; Thompson et al., 2009). Given that SES-related disparities in brain connectivity may stem from a combination of prenatal exposures, postnatal environments, and genetic predispositions, the observed negative loading may capture broader socioeconomic influences rather than a direct teratogenic effect of prenatal substance exposure. However, disentangling these effects is challenging, as the precise mechanisms through which prenatal exposures contribute to structural brain network alterations remain unclear. Future studies leveraging genome-environment interaction analyses, longitudinal neuroimaging, and causal inference approaches (e.g., Mendelian randomization) are essential to elucidate how prenatal risk factors, SES, and genetic predispositions collectively shape neurodevelopmental outcomes.

Phenotype mode 2 presents a complex pattern, linking standing height, perinatal risk factors, and sensorimotor brain networks. The lower network efficiency in the precentral and postcentral gyri, superior frontal gyrus, and thalamus suggests a potential connection to early neurodevelopmental processes, particularly those involved in motor and sensory integration. Given the variability in loading estimates, further research is needed to determine whether these associations reflect specific neurodevelopmental mechanisms or statistical artifacts.

Several limitations should be acknowledged. First, the cross-sectional design precludes causal inferences, necessitating longitudinal research to validate the mediating role of brain networks in shaping developmental trajectories of cognitive and mental health. Second, gene-by-environment (G×E) interactions were not explicitly modeled, limiting our ability to assess whether socioeconomic status (SES) moderates genetic influences on brain connectivity. Future research should integrate G×E interaction analyses to better understand how genetic predispositions interact with environmental contexts in shaping neurodevelopment. Finally, although sparse canonical correlation analysis (SCCA) provided a powerful multivariate approach, it assumes linear relationships between brain networks and genetic/environmental factors, which may oversimplify complex neurodevelopmental processes. Future studies should consider non-linear modeling approaches and more sophisticated causal inference methods (e.g., Mendelian randomization, structural equation modeling) to capture the intricate interplay of genes, environments, and brain development.

Addressing these gaps will enhance the robustness of future research, ultimately contributing to a more comprehensive understanding of how genetic and environmental factors shape brain development. These insights may inform early intervention strategies aimed at mitigating neurodevelopmental disparities, such as targeted cognitive training, socioeconomic policy reforms, or school-based enrichment programs that support children from disadvantaged backgrounds.

## Ethics Statement

This study analyzed data from the Adolescent Brain Cognitive Development (ABCD) study. All participants provided informed assent, and their parents or legal guardians provided informed consent before participating in the ABCD study.

## Data Availability

All original data are publicly available from the NDA (https://nda.nih.gov/abcd/). Mook data, which corresponds to the processed data used in this study, was generated from conditional GAN for tabular data (Xu et al., 2019) and are available from this site (https://github.com/Transconnectome/ABCD-brain-network-SCCA).

## Acknowledgments

This work was supported by the National Research Foundation of Korea(NRF) grant funded by the Korea government(MSIT) (No. 2021R1C1C1006503, RS-2023-00266787, RS-2023-00265406, RS-2024-00421268), by Creative-Pioneering Researchers Program through Seoul National University(No. 200-20240057), by Semi-Supervised Learning Research Grant by SAMSUNG(No.A0426-20220118), by Identify the network of brain preparation steps for concentration Research Grant by LooxidLabs(No.339-20230001), by Institute of Information & communications Technology Planning & Evaluation (IITP) grant funded by the Korea government(MSIT) [NO.RS-2021-II211343, Artificial Intelligence Graduate School Program (Seoul National University)] by the MSIT(Ministry of Science, ICT), Korea, under the Global Research Support Program in the Digital Field program(RS-2024-00421268) supervised by the IITP(Institute for Information & Communications Technology Planning & Evaluation), by the National Supercomputing Center with supercomputing resources including technical support(KSC-2023-CRE-0568) and by the Ministry of Education of the Republic of Korea and the National Research Foundation of Korea (NRF-2021S1A3A2A02090597), and by Artificial intelligence industrial convergence cluster development project funded by the Ministry of Science and ICT(MSIT, Korea) & Gwangju Metropolitan City.

## Data Availability

All original data are publicly available from the NDA (https://nda.nih.gov/abcd/). For rapid replication, we provide synthetic data, whose distributions were matched to the original data, generated using conditional GAN for tabular data (Xu et al., 2019) (https://github.com/Transconnectome/ABCD-brain-network-SCCA).

## Code Availability

Code is available from here: https://github.com/Transconnectome/ABCD-brain-network-SCCA.

## Author contributions

- Jungwoo Seo, Conceptualization, Data curation, Formal analysis, Investigation, Methodology, Project administration, Software, Validation, Visualization, Writing – original draft, Writing – review and editing
- Eunji Lee, Data curation, Writing – original draft, Writing – review and editing
- Bo-Gyeom Kim, Data curation, Writing – review and editing
- Gakyung Kim, Data curation, Writing – original draft, Writing – review and editing
- Yoonjung Yoonie Joo, Data curation, Writing – review and editing
- Jiook Cha, Conceptualization, Supervision, Project administration, Funding acquisition, Resources, Data curation, Writing – original draft, Writing – review and editing.

## Supplementary Materials

### Details on Genotype Data

The saliva DNA samples of study participants were collected, and 733,293 single nucleotide polymorphisms (SNPs) were genotyped at Rutgers University Cell and DNA Repository (RUCDR) with Affymetrix NIDA Smoke Screen Array. Using PLINK 1.90, we excluded SNPs with genotype call rate <95%, sample call rate <95%, and minor allele frequency (MAF) <1% before imputation. The genotypes were imputed using the Michigan Imputation Server (Das et al., 2016) using the 1000 Genome phase3 version5 panel (Genomes Project et al., 2015) with Eagle v2.4 phasing (Loh et al., 2016). Then, the imputed variants with INFO score > .3 that did not meet our quality control criteria (i.e., call rate <95%, MAF <1%, and Hardy– Weinberg equilibrium p-value <1e-6) were additionally filtered out. To address potential bias derived from genetically diverse and related family members in the ABCD study, we employed PC-Air (Conomos et al., 2015) and PC-Relate (Conomos et al., 2016) to obtain genetically unrelated individuals beyond 4th-degree relatives (i.e., kinship coefficient >0.022) and to remove outliers beyond 6 SD limits from the center of ancestrally informative principal component (PC) space. After quality control procedures, we included a total of 11,301,999 variants in 10,199 multiethnic participants. From these, we excluded first-, second-, or third-degree related samples, resulting in 8,620 unrelated multiethnic participants, among whom 6,555 were of European ancestry.

### Details on Polygenic Scores (PGSs)

Polygenic scores (PGSs) analyzed in this study are the same PGSs data used in (Joo et al., 2024). PGSs were derived using summary statistics from publicly available genome-wide association studies (GWAS). Thirty traits were selected based on relevance to cognitive, psychiatric, and behavioral outcomes. The chosen GWAS include attention-deficit/hyperactivity disorder (ADHD) (Demontis et al., 2019), cognitive performance (CP) (Lee et al., 2018), educational attainment (EA) (Lee et al., 2018), major depressive disorder (MDD) (Wray et al., 2018), insomnia (Jansen et al., 2019), snoring (Jansen et al., 2019), intelligence quotient (IQ) (Savage et al., 2018), post-traumatic stress disorder (PTSD) (Nievergelt et al., 2019), depression (DEP) (Howard et al., 2019; Shen et al., 2020), body mass index (BMI) (Akiyama et al., 2017; Locke et al., 2015), alcohol dependence (ALCDEP) (Walters et al., 2018), autism spectrum disorder (ASD) (Grove et al., 2019), automobile speeding propensity (ASP) (Akiyama et al., 2017), bipolar disorder (BIP) (Stahl et al., 2019), cannabis during lifetimes (Cannabis) (Pasman et al., 2019), ever smoker (Karlsson Linner et al., 2019), shared effects on five major psychiatric disorder (CROSS) (Cross-Disorder Group of the Psychiatric Genomics, 2013), alcoholic drinks consumption per week (Drinking) (Karlsson Linner et al., 2019), anorexia nervosa (Watson et al., 2019), neuroticism (Nagel et al., 2018), obsessive-compulsive disorder (OCD) (International Obsessive Compulsive Disorder Foundation Genetics & Studies, 2018), first principal components of four risky behaviors (Risky Behav) (Karlsson Linner et al., 2019), general risk tolerance (RiskTol) (Karlsson Linner et al., 2019), schizophrenia (SCZ) (Bipolar et al., 2018; Lam et al., 2019), worrying (Nagel et al., 2018), anxiety (Otowa et al., 2016), subjective well-being (SWB) (Okbay et al., 2016), general happiness (UK Biobank GWAS. Neale Lab. http://www.nealelab.is/ukbiobank/), and general happiness for health (happiness-health) (UK Biobank GWAS. Neale Lab. http://www.nealelab.is/ukbiobank/) and meaningful life (happiness-meaning) (UK Biobank GWAS. Neale Lab. http://www.nealelab.is/ukbiobank/).

For 25 of these traits, where only European-based GWAS were available, we calculated polygenic scores using European-based GWAS summary statistics. For five traits where multiethnic GWAS results were available (PTSD, DEP, BMI, ALCDEP, SCZ), we calculated polygenic scores using both European-based and multiethnic-based GWAS summary statistics. In the European-only analysis, we used polygenic scores for all 30 traits calculated based on European-based GWAS.

The GWAS summary statistics were used as input for PRS-CS (Ge et al., 2019), a Bayesian regression method, to estimate the posterior effect sizes of SNPs. The final scores were calculated using PLINK v1.9. To optimize the scores, we followed the suggestion of the original PRS-CS paper and chose the optimal global shrinkage hyperparameter (phi, φ) from among four possible values: 1, 1e-2, 1e-4, and 1e-6. The validation procedure was carried out within 14 PGSs (i.e., DEP, MDD, ADHD, general happiness, happiness-health, happiness-meaning, SWB, insomnia, snoring, BMI, PTSD, CP, EA, IQ) that had related measures in the ABCD study. For each PGS, we performed linear regression of the phenotype variable with each of the four scores and covariates (sex, age, and the first ten genetic PCs), and then, based on *R^2^* and beta coefficient of PGS, selected one of the four PGSs. The remaining 16 PGSs was automatically validated by PRS-CS-auto (Ge et al., 2019), which select the optimal value of global shrinkage parameter employing a Bayesian approach. Finally, to minimize the bias from population stratification, we residualized the final PGSs with the first ten genetic PCs.

**Supplementary Table 1.** Demographic information of the supplementary analysis participants (multiethnic analysis and complete data samples).

|  |  | PGS – BNM<br>(n=7,297) |  | ENV – BNM<br>(n=4,215) |  | Pheno – BNM<br>(n=6,598) |  |
| --- | --- | --- | --- | --- | --- | --- | --- |
|  |  | train | test | train | test | train | test |
| N | total | 5,837 | 1,460 | 3,372 | 843 | 5,278 | 1,320 |
| Sex | Male | 3,087 | 765 | 1,790 | 440 | 3,031 | 758 |
|  | Female | 2,747 | 695 | 1,581 | 403 | 2,247 | 562 |
|  | Other | 3 | 0 | 1 | 0 | 0 | 0 |
| Race | White | 3,251 | 780 | 1,702 | 451 | 3,036 | 765 |
|  | Black | 798 | 207 | 474 | 103 | 674 | 161 |
|  | Hispanic | 1,176 | 302 | 781 | 183 | 933 | 228 |
|  | Asian | 18 | 6 | 86 | 21 | 94 | 19 |
|  | Other | 594 | 165 | 329 | 85 | 541 | 147 |

**Supplementary Figure 1.**
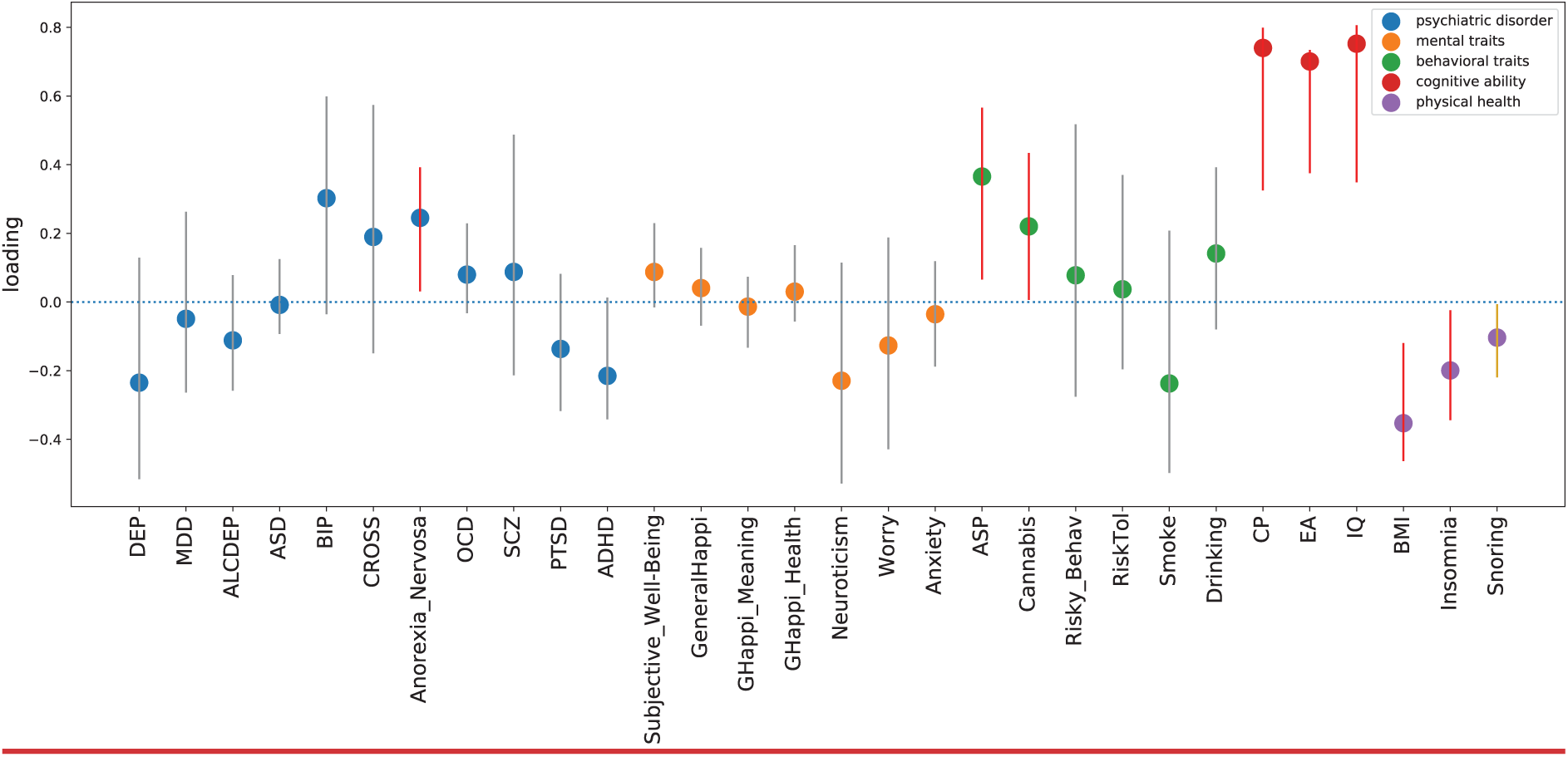
SCCA Loadings of Polygenic Scores in European Samples.

**Supplementary Figure 2.**
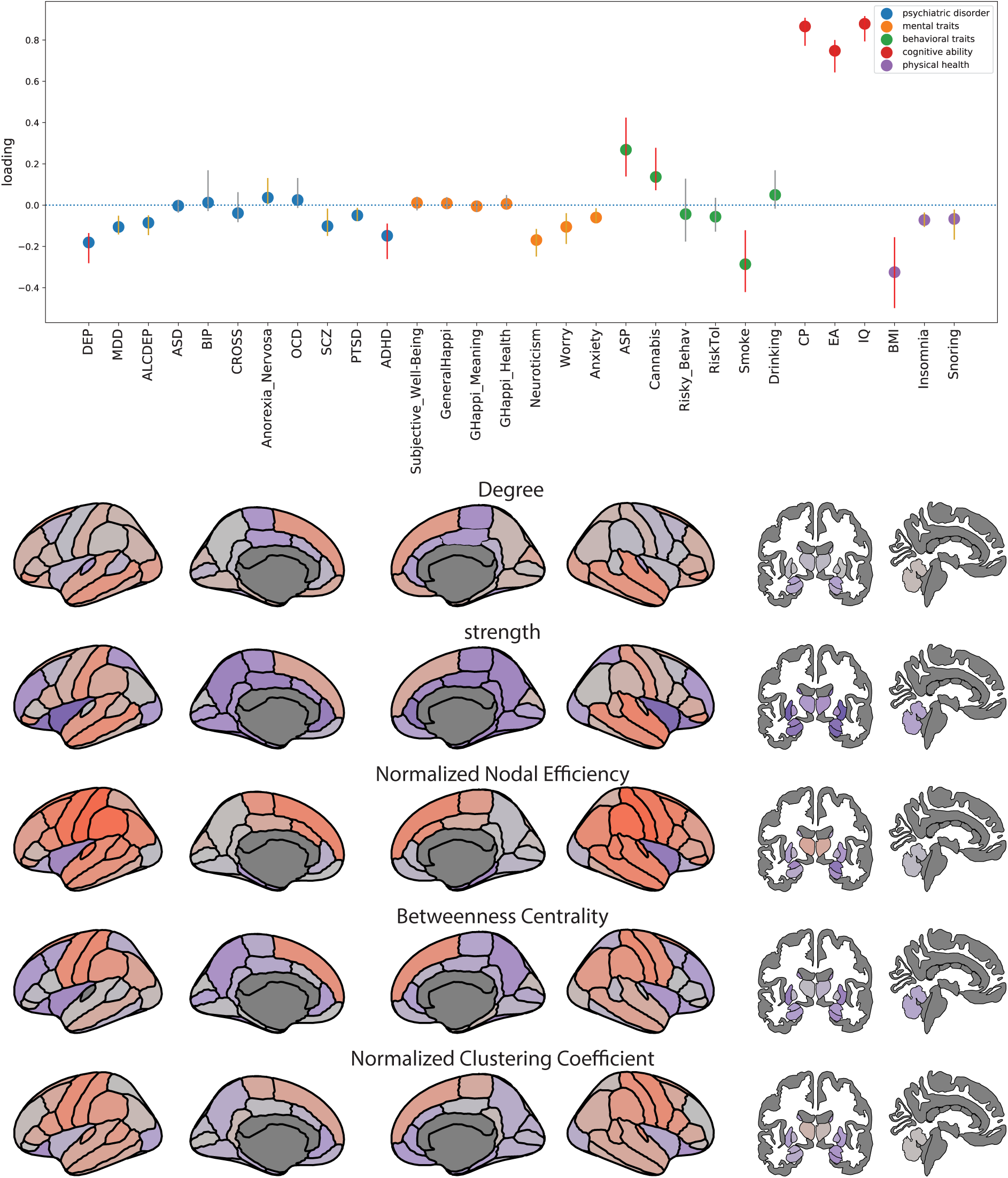
SCCA Loadings of Polygenic Scores and Brain Network Measures in Multiethnic Samples.

**Supplementary Figure 3.**
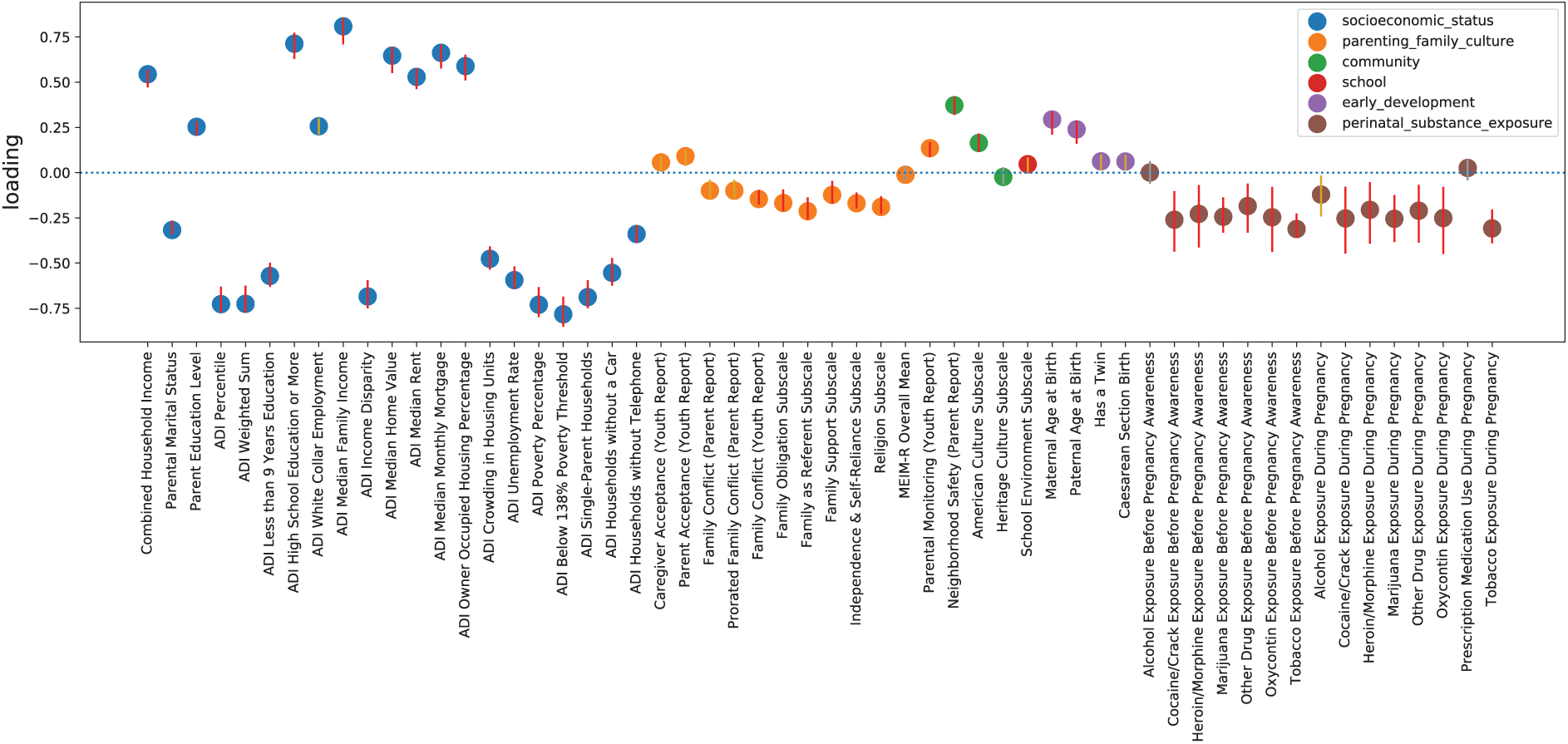
SCCA Loadings of Environmental Variables in the Main Analysis.

**Supplementary Figure 4.**
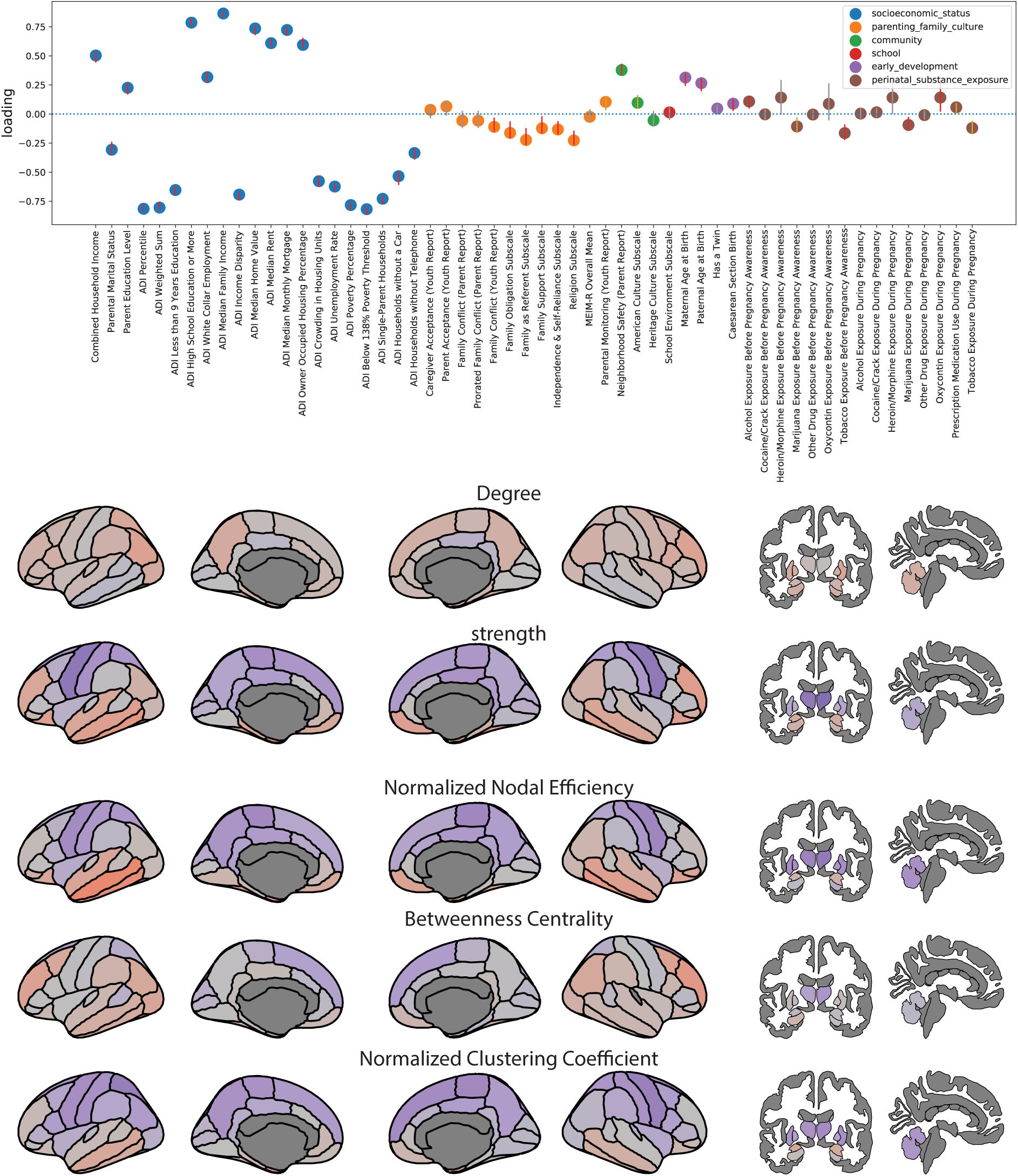
SCCA Loadings of Environmental Variables and Brain Network Measures in Non-Imputed Samples.

**Supplementary Figure 5.**
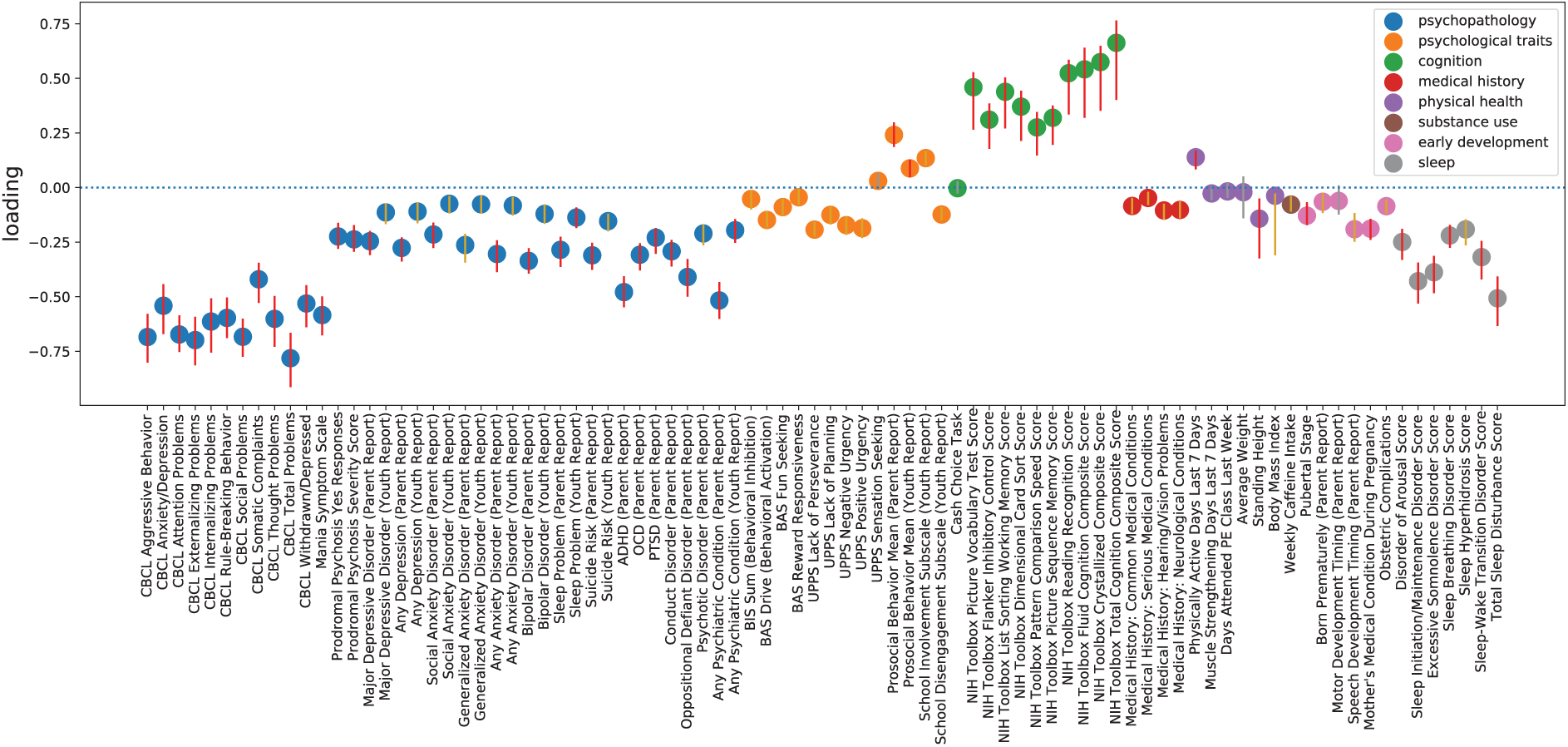
SCCA Loadings of Phenotype Variables in the Main Analysis Mode 1.

**Supplementary Figure 6.**
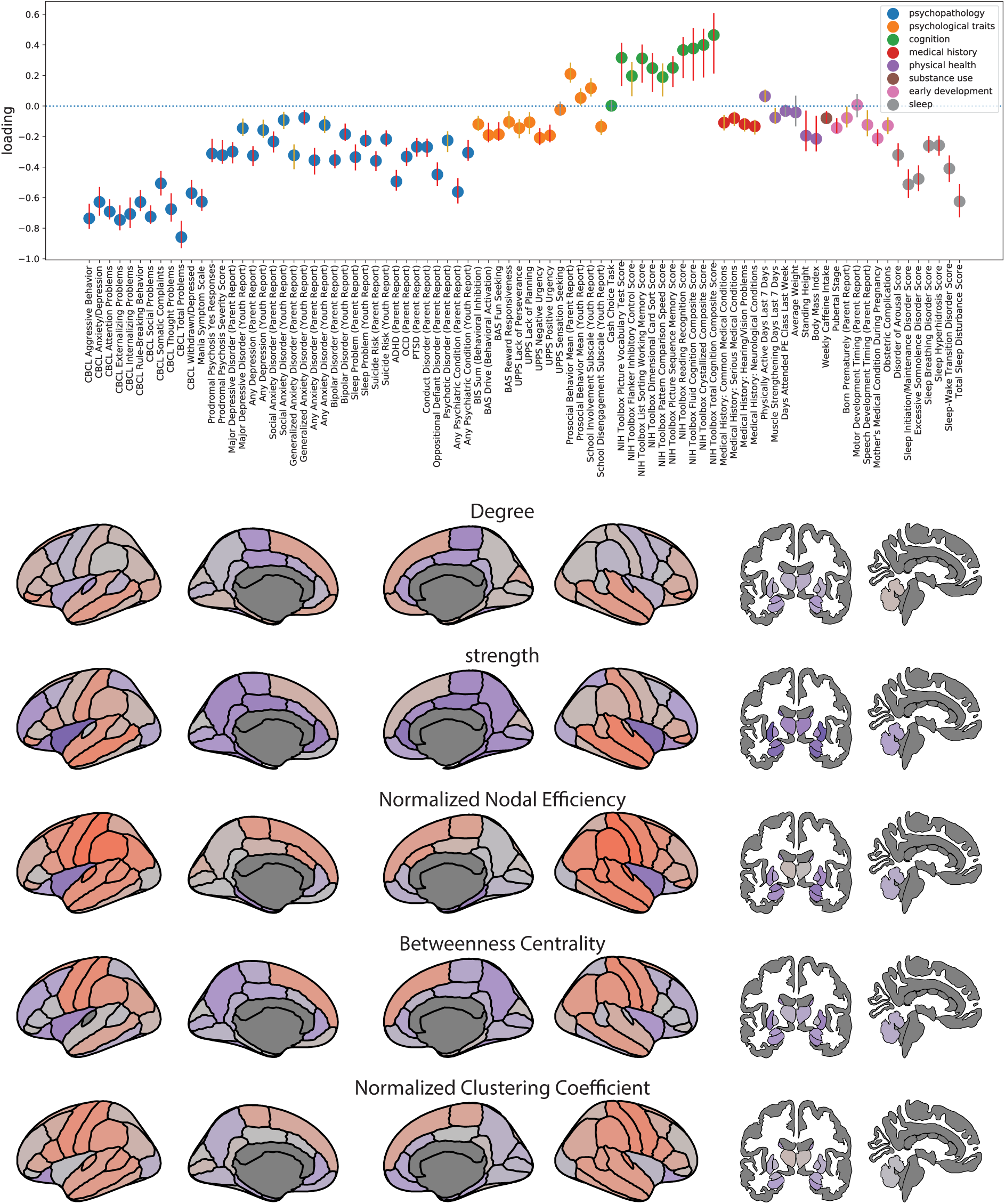
SCCA Loadings of Phenotype Variables and Brain Network Measures in Non-Imputed Samples Mode 1.

**Supplementary Figure 7.**
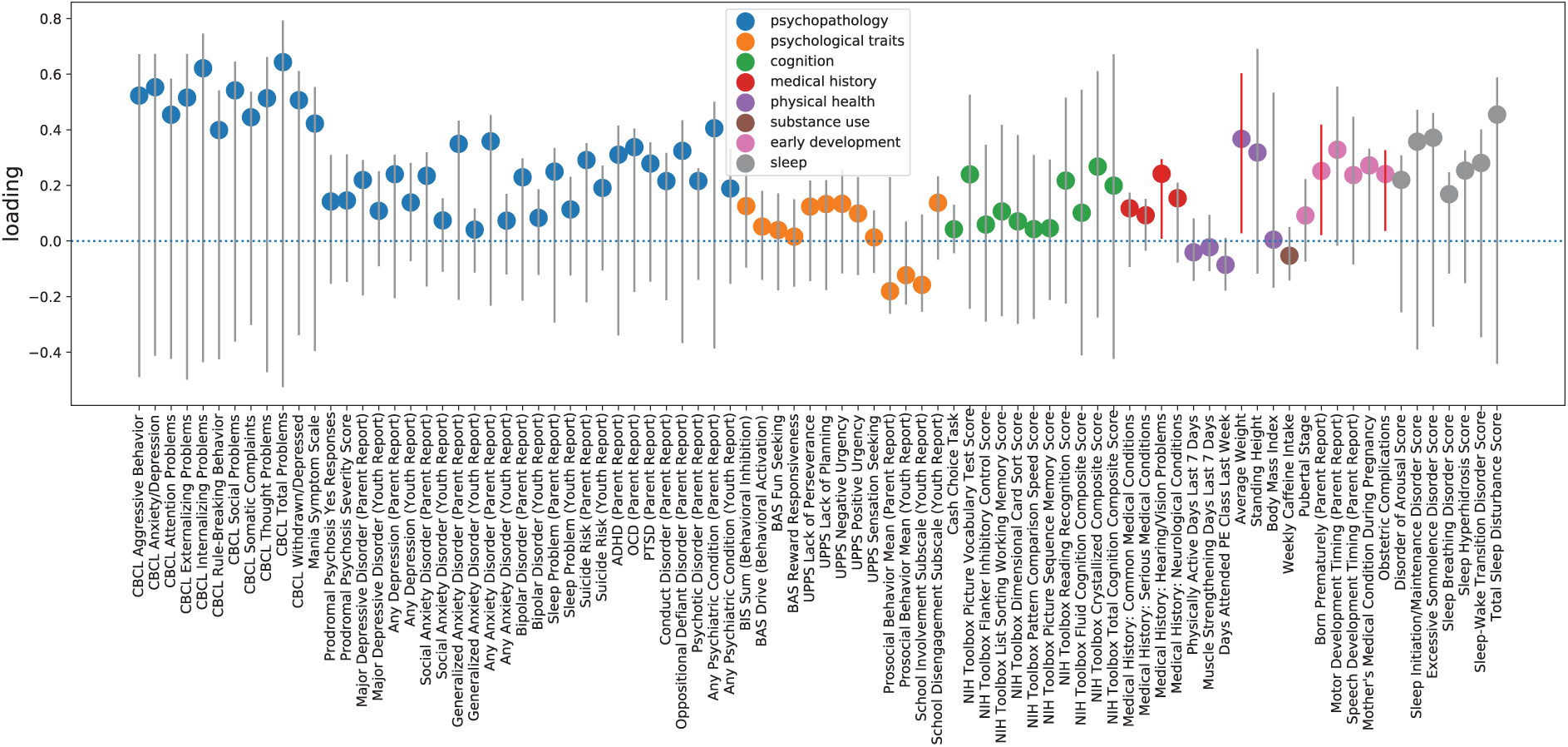
SCCA Loadings of Phenotype Variables in the Main Analysis Mode 2.

**Supplementary Figure 8.**
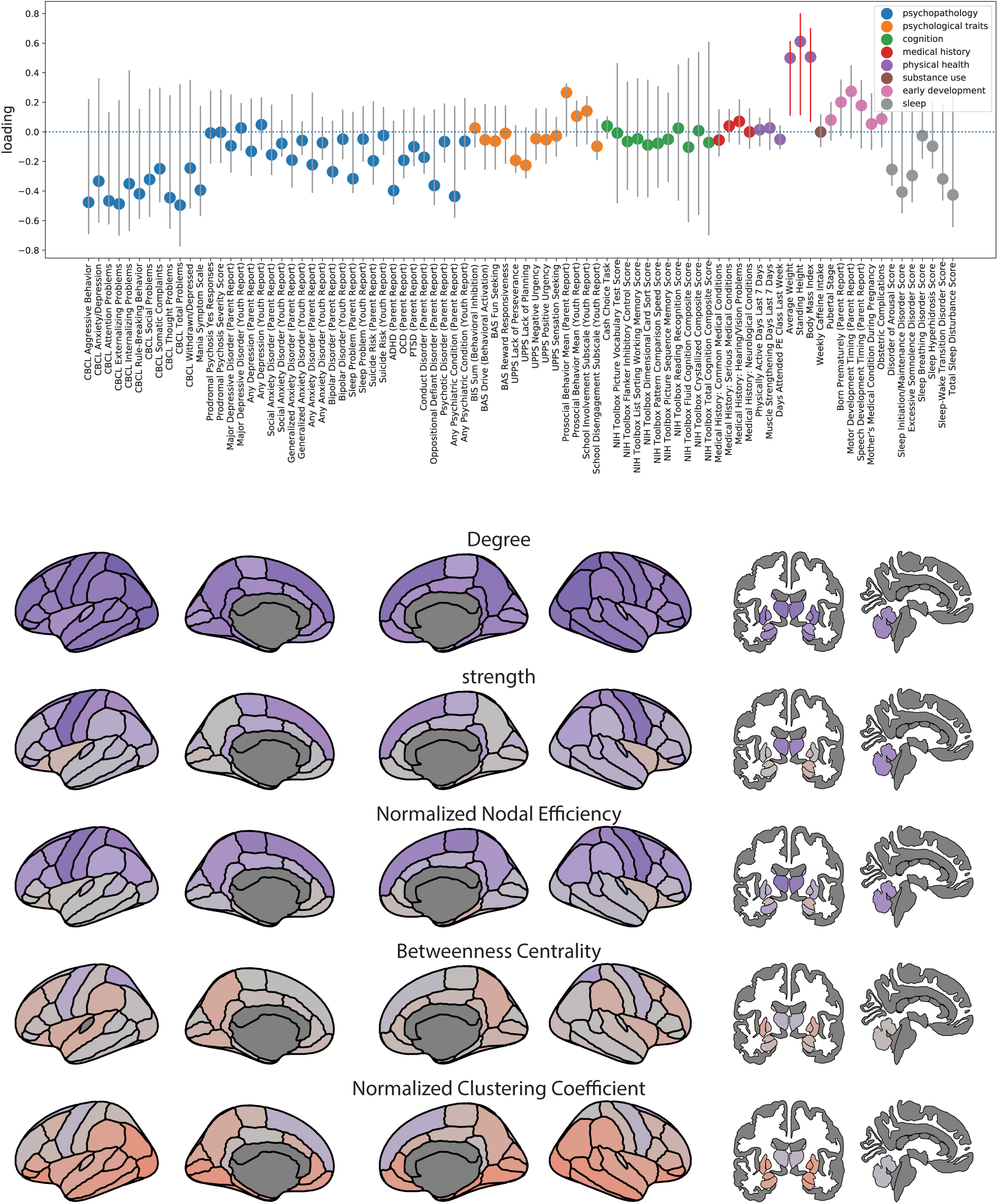
SCCA Loadings of Phenotype Variables and Brain Network Measures in Non-Imputed Samples Mode 2.

